# Global burden, trends, and projections of pelvic organ prolapse in postmenopausal elderly women from 1990 to 2021: a systematic analysis based on GBD and Mendelian randomization

**DOI:** 10.1101/2025.08.28.25334618

**Authors:** Zhengkun Wang, Qiwei Wang, Yi Rong, Yikai Wang, Weibing Shuang

## Abstract

**Background:** Pelvic organ prolapse (POP) is a widespread disease in women; however, the global burden and trend of POP in postmenopausal women (PMW) remain poorly characterized.

**Methods:** Based on the Global Burden of Disease (GBD) 2021 database and genome-wide association studies (GWAS), this study assessed the burden and risk factors of POP in PMW worldwide. We employed the slope index of inequality (SII) and concentration index (CI) to evaluate regional health inequalities. Additionally, age-period-cohort (APC) analysis was integrated with the Bayesian age-period-cohort (BAPC) model to examine temporal trends and project future disease burden trends. Furthermore, we conducted a two-sample Mendelian randomization (MR) analysis to evaluate the causal relationships between risk factors and POP.

**Results:** From 1990 to 2021, the global incidence, prevalence, and disability-adjusted life years (DALYs) of POP in PMW increased by 92.96%, 85.21%, and 83.78%, respectively. However, in 2021, the global age-standardized disability rate (ASDR) of POP was 16.21% lower than in 1990. Notably, regions with a low sociodemographic index (SDI) experienced statistically significant increases in disease burden. Additionally, individuals with obesity faced a higher risk of POP, while those with higher education levels had a lower risk. According to the BAPC model, global cases of POP among PMW are projected to reach 114.67 million by 2050, while the age-standardized incidence rate (ASIR) is expected to decline by only 3.49%.

**Conclusion:** The global burden of POP among PMW has increased in total cases, despite declining age-standardized rates. Aging populations are projected to further exacerbate this burden, underscoring the need for targeted strategies to address high-risk populations.

## Introduction

Pelvic organ prolapse (POP) is a common manifestation of dysfunction in the female pelvic floor support system. It is characterized by the abnormal descent of pelvic organs, such as the uterus, bladder, and rectum, caused by structural damage to the pelvic ligaments, fascia, and muscles. The primary clinical features include vaginal wall protrusion and uterine prolapse, which are often accompanied by intestinal and urinary dysfunction ^[1]^.

Menopause, marking the end of a woman’s reproductive age, is characterized by a significant decline in estrogen levels, which leads to the degeneration of pelvic floor support structures ^[2]^. Epidemiological data from the Global Burden of Disease (GBD) database indicate that POP affected approximately 127.68 million individuals worldwide in 2021. Of these, postmenopausal women (PMW) (55+ years) comprised 76.44 million cases, accounting for 61.46%. Research has established that aging represents the most significant risk factor for POP, followed by parity and obesity. Epidemiological studies indicate that women aged 70–79 are most likely to seek medical attention for symptomatic POP ^[3]^. In the United States, about 40% of POP patients are over 65 years old ^[4]^. Due to the global trend of population aging, the average life expectancy has increased from 65.42 years in 1990 to 73.52 years in 2019 ^[5]^. By 2050, projections indicate that the population aged over 60 will double to 2.1 billion, with the number of individuals over 80 reaching 426 million ^[6]^. As a result, the number of postmenopausal elderly women affected by POP is projected to rise significantly.

The GBD study provides the most comprehensive epidemiological assessment of POP to date ^[7]^. Previous research primarily focused on females across all age groups, and thus the global burden and temporal trends of POP in PMW have remained poorly characterized. Using the latest GBD 2021 data, this study systematically quantifies the burden of POP among PMW across 204 countries and territories, performing granular stratified analyses based on the Socio-demographic Index (SDI).

## Materials and methods

### Data sources

The data are derived from the GBD 2021 database (https://vizhub.healthdata.org/gbd-results/, accessed on June 23, 2025) and the MRC Integrative Epidemiology Unit (IEU) OpenGWAS project (https://gwas.mrcieu.ac.uk/, accessed on June 26, 2025).

The study analyzed indicators associated with POP, including incidence, prevalence, and Disability-Adjusted Life Years (DALYs), across all countries and territories from 1990 to 2021. The data were stratified by location, SDI, and age group to examine temporal and spatial patterns in the disease burden among PMW aged 55 and older. DALYs are used as a metric to measure the total health loss caused by specific diseases or risk factors in population health. The SDI quantifies the socio-economic status of a country or location, reflecting its association with health outcomes. The SDI scale ranges from 0 (indicating high fertility, low education, and low income) to 1 (low fertility, high education, and high income) ^[8]^.

Since the GBD database is publicly accessible and contains no personally identifiable or sensitive data, and all genome-wide association studies (GWAS) included in this Mendelian Randomization (MR) study were approved by their respective institutional review boards, this study was exempt from institutional ethical review.

### Statistical analysis

POP primarily affects the quality of life of patients and is rarely associated with direct mortality. The crude rates (prevalence, incidence, and DALYs) obtained in this study provide fundamental metrics to measure the epidemiological trends of diseases; however, differences in population age distribution may lead to variations in the burden of POP. To enhance the consistency of statistical measures, the crude rates were adjusted by applying different weights based on the age composition of the population, resulting in the age-standardized rate (ASR). To reflect the annual trends in the global burden of POP from 1990 to 2021, we calculated the Estimated Annual Percentage Change (EAPC) for the ASIR, ASPR, and ASDR. The rates are expressed as the mean estimated value per 100,000 population, along with a 95% uncertainty interval (UI).

By analyzing data from 1990 to 2021, this study quantifies changes in each index and systematically examines the evolution of epidemiological characteristics related to POP among PMW across multidimensional classifications, including region, age and year, through descriptive analysis and visual presentation. All statistical analyses were performed using R software (version 4.3.3, http://www.r-project.org/).

### Health inequality analysis

In this study, health inequalities were analyzed using DALYs and ASDR. Absolute and relative income-related inequalities were assessed using the Slope Index of Inequality (SII) and the Concentration Index (CI). The SII was calculated using a weighted regression model ranking ASDR and SDI nationally, while CI was derived from a Lorenz concentration curve, plotting cumulative population ratio against cumulative SDI-related DALYs, and was computed through area integration. Negative values for both the SII and CI indicate that a lower SDI corresponds to a higher disease burden; larger absolute magnitudes signify greater inequality ^[9]^.

### Frontier analysis

To investigate the relationship between POP disease burden and the SDI, this study applied frontier analysis using the GBD database. The analysis aimed to identify opportunities for reducing disease burden across various countries and territories through SDI assessment ^[10]^. The frontier reflects the theoretical minimum burden achievable at a given SDI level, while the effective difference—defined as the deviation from this frontier—quantifies the gap between observed and optimal burden levels. Countries closer to the frontier demonstrate efficient disease control, while those further from the frontier highlight substantial potential for improvement.

### Age-Period-Cohort analysis

This study employed an Age-Period-Cohort (APC) model to analyze the data, treating age, period, and cohort as independent variables. Within this framework, the age effect represents variations in outcome risk across different age groups. The period effect quantifies temporal changes that concurrently affect all age groups. The cohort effect reflects differences in outcome risk among individuals born in the same year. To ensure smooth time-effect curves and reduce the number of parameters, age-specific incidence rates were aggregated into 5-year blocks (55–59, 60–64, …, 95+ years). Consistent with APC modeling requirements for equal interval categorization, calendar periods were similarly grouped into contiguous 5-year blocks (1992–1996 through 2017–2021). Key estimable functions included: (1) Net drift, representing the overall annual percentage change in incidence rates; (2) Local drifts, detailing age-specific annual percentage changes across periods and cohorts; (3) Longitudinal age curves, depicting fitted age-specific rates over time within the referent cohort (adjusted for period deviations); and (4) Period/cohort rate ratios (RRs), indicating age-specific rate ratios relative to reference periods or cohorts. An RR > 1 indicates an elevated incidence rate, while an RR < 1 indicates a reduced incidence rate ^[11]^.

### Bayesian age-period-cohort analysis

We employed the Bayesian age-period-cohort (BAPC) model to project the future burden of POP among PMW. This model, accessible through the BAPC R package, integrates age, period, and cohort effects within a comprehensive Bayesian framework. For Bayesian inference, we utilized the Integrated Nested Laplace Approximation (INLA) method. INLA efficiently approximates the marginal posterior distributions of model parameters, thereby providing a computationally efficient alternative to conventional Markov Chain Monte Carlo (MCMC) methods.

### Mendelian randomization analysis

The Mendelian randomization (MR) method evaluates causal relationships between environmental exposures and outcomes using genetic variants from GWAS as instrumental variables (IVs). The datasets were obtained from the MRC Integrative Epidemiology Unit (IEU) OpenGWAS project (https://gwas.mrcieu.ac.uk/). The significance threshold was set at P < 5*10^-8^ to satisfy the instrument strength assumption. Single-nucleotide polymorphisms (SNPs) in linkage disequilibrium (LD) (r^2^ < 0.001) were excluded to ensure genetic independence. All selected SNPs had F-statistics > 10, indicating sufficient instrumental variable strength. The inverse-variance weighted (IVW) method served as the primary approach, supplemented by MR-Egger regression and weighted median methods for sensitivity analyses.

## Results

### Global POP burden from 1990 to 2021

From 1990 to 2021, the global number of POP incidence among PMW increased from 3.33 million to 6.44 million, representing a 92.96% increase. Similarly, the number of prevalent cases rose from 41.27 million to 76.44 million , marking a 85.21% increase. Additionally, DALYs increased from 0.13 million to 0.24 million, reflecting a 83.78% increase. Conversely, the global ASIR decreased from 927.64 to 816.74 per 100,000 population in 2021, EAPC = -0.256. Similarly, the ASPR declined from 11499.41 to 9712.94 per 100,000 population, EAPC = -0.373. The ASDR per 100,000 population declined from 36.03 to 30.19, EAPC = -0.391 (Table 1 and S1 Table).

**Table 1.**
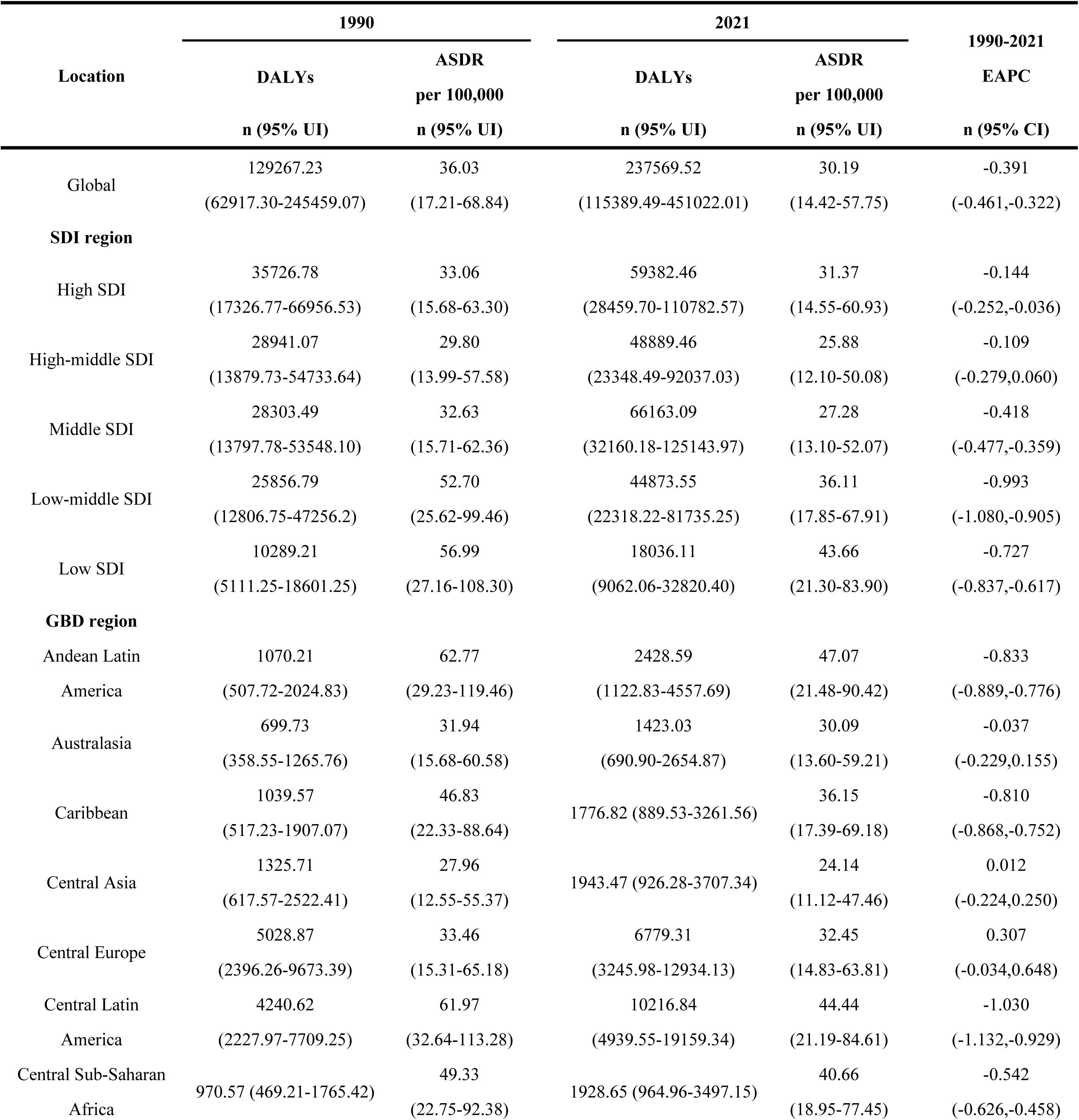
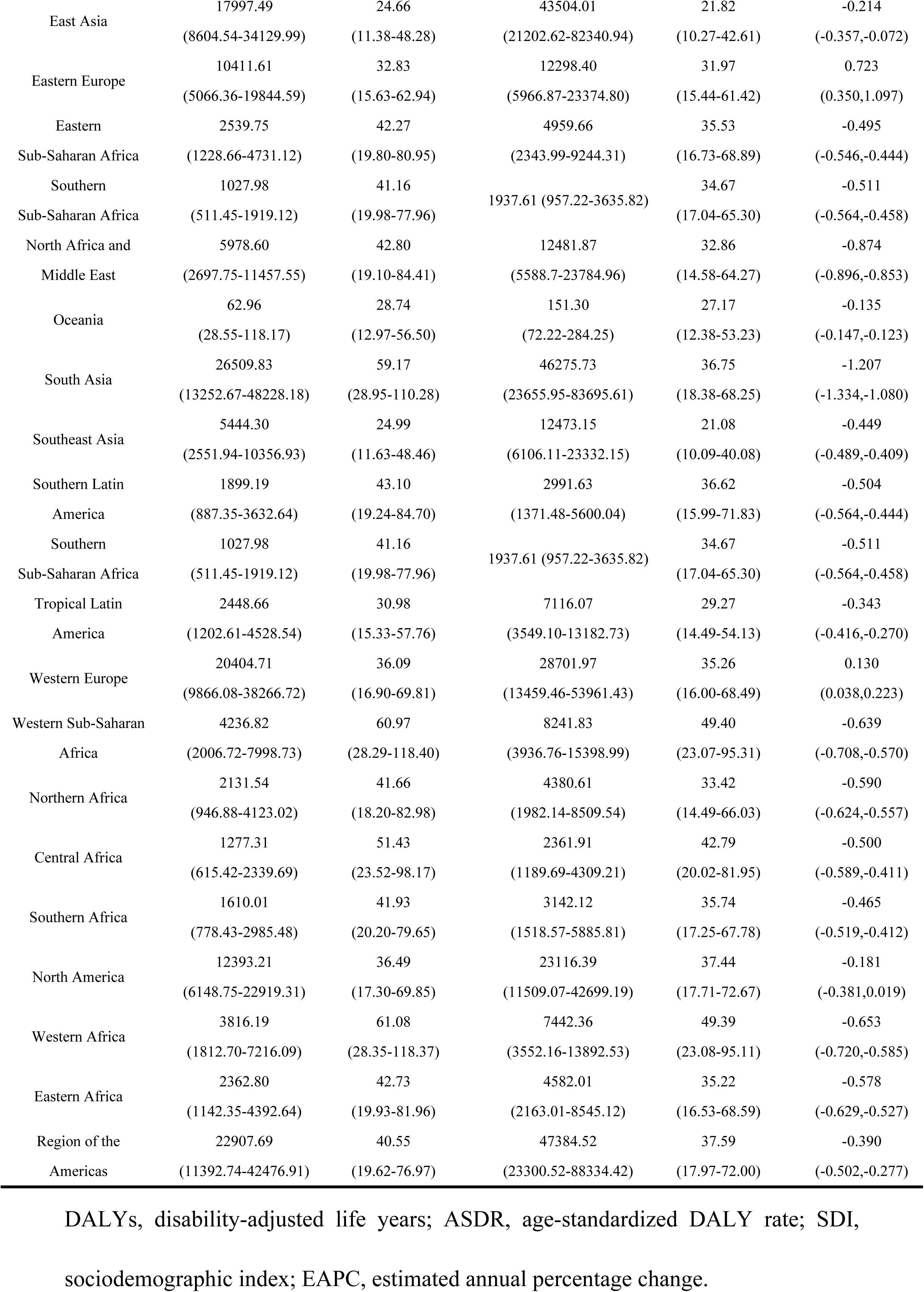
Age-standardized DALYs rates and corresponding EAPC of POP in postmenopausal women globally and regions in 1990 and 2021.

### POP burden in SDI regions

Analysis stratified by SDI level demonstrates that the overall burden of POP among PMW declined from 1990 to 2021. In 2021, the highest burden of disease was observed in the Low SDI region, with ASIR, ASPR, and ASDR rates of 1000.79, 13723.76, and 43.66 per 100,000 population respectively. Conversely, the lowest burden was found in the High-middle SDI region, with corresponding rates of 738.80, 8434.79, and 25.88 per 100,000 population. The most rapid declines in EAPC occurred in the Low-middle SDI region, with values of -0.814 (ASIR), -1.032 (ASPR), and -0.993 (ASDR). Notably, the High SDI region showed an increasing trend in ASIR (EAPC = 0.038) . Furthermore, compared to other SDI regions, High SDI exhibited the slowest declines in ASPR (EAPC = -0.069) (S1 Table).

Analysis of GBD data from 1990 to 2021 demonstrates a significant negative correlation (R = -0.57, 95% CI: -0.61 to -0.51, p < 0.001) between the SDI and the ASDR of POP in PMW. The ASDR is relatively higher in low-SDI regions. As the SDI increased to approximately 0.5, the ASDR exhibited a transient increase, followed by a continuous decline. In high-SDI regions, the ASDR is relatively lower, according to the chart findings (Fig 1 and S1 Fig).

**Fig 1.**
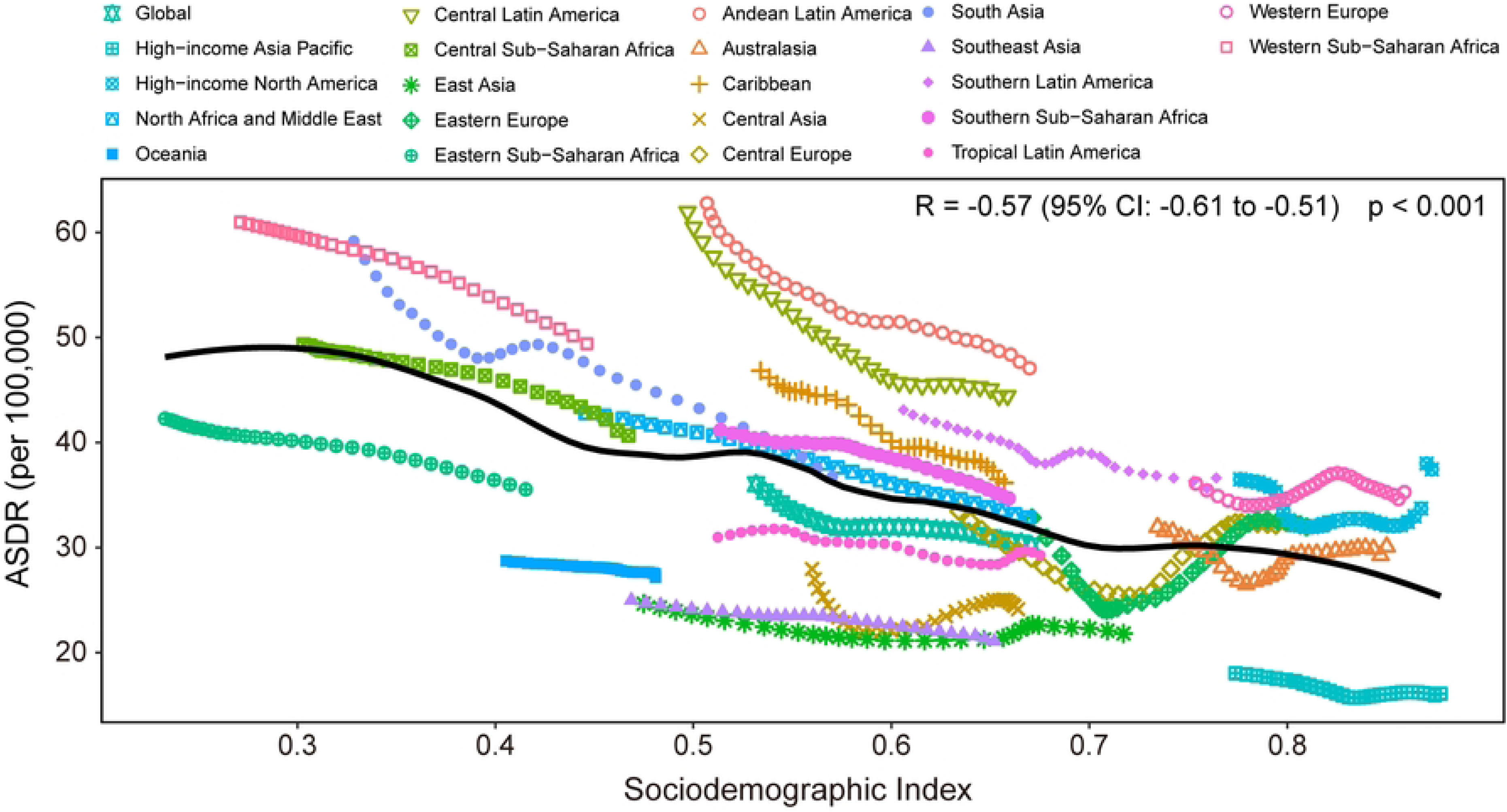
Age-standardized DALY rate of POP in postmenopausal women for the 21 GBD regions by SDI, 1990-2021. ASDR, age-standardized DALY rate; SDI, sociodemographic index.

### POP burden in all locations

According to GBD 2021 data, the highest ASIR, ASPR, and ASDR for POP among PMW were observed in Western Sub-Saharan Africa, Andean Latin America, and Central Latin America. Specifically, Western Sub-Saharan Africa had the highest ASIR (1,172.23 per 100,000), while the lowest ASIR was observed in High-income Asia Pacific (582.53 per 100,000). Between 1990 and 2021, ASIR increased in only 11 GBD regions, with Eastern Europe showing the most significant rise (EAPC=0.669). This region also demonstrated the fastest growth in ASPR (EAPC=0.739) and ASDR (EAPC=0.723) (Fig 2 and S1 Table).

**Fig 2.**
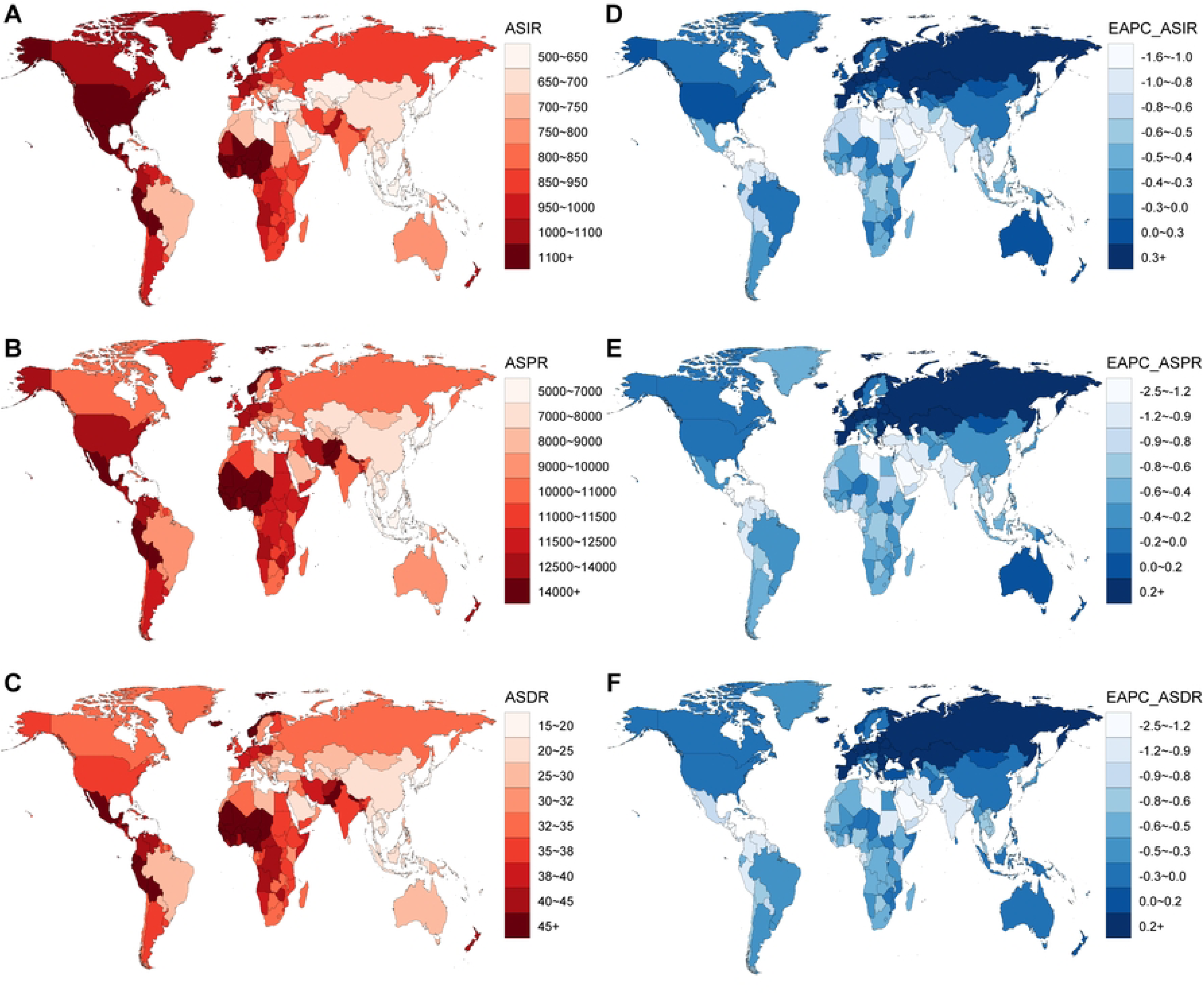
Global POP burden in 204 countries and territories. (A-C) The ASIR, ASPR, and ASDR in 2021; (D-F) The EAPC of ASIR, ASPR, and ASDR between 1990 and 2021. DALYs, disability-adjusted life years; ASIR, age-standardized incidence rate; ASPR, age-standardized prevalence rate; ASDR, age-standardized DALY rate; EAPC, estimated annual percentage change.

In 2021, the location with the highest ASPR and ASDR among PMW was Niger, the respective rates were 20,227.18 and 62.77 per 100,000. In contrast, Japan had the lowest ASPR and ASDR of POP globally, with rates of 5039.70 and 15.59 per 100,000, respectively. The three countries with the highest national ASIR of POP were Niger (1333.55 per 100,000), Chad (1276.53 per 100,000), and Mali (1258.26 per 100,000). In contrast, Thailand (536.29 per 100,000), Azerbaijan (553.52 per 100,000), and Armenia (554.08 per 100,000) had the lowest ASIR. Compared with 1990, the EAPC for ASIR, ASPR and ASDR increased in approximately 31 countries among PMW. Among them, Russian Federation had the highest EAPC increase: 0.829 (ASIR), 0.945 (ASPR), and 0.923 (ASDR); Nepal showed the most significant decline: -1.577 (ASIR), -2.382 (ASPR), and -2.328 (ASDR) (Fig 2 and S1 Table).

### Health inequality of POP burden

Based on the analysis of global health inequality models for POP among PMW from 1990 to 2021, the SII for DALYs showed a reduction in its absolute magnitude. The SII value increased (became less negative) from -20.66 in 1990 to -7.98 in 2021, representing a 58.30% decrease in absolute inequality magnitude. Lorenz curve analysis revealed that the CI increased (became less negative) from -0.07 to -0.03. Both SII and CI values are negative, indicating a disproportionate concentration of disease burden in low-SDI regions. The reduction in the absolute magnitude of these negative values demonstrates improvements in both absolute and relative inequality levels between the highest and lowest SDI groups. For example, China and India, as the most populous nations, both exhibited declining crude DALY rates; however, India’s crude DALY rate remained notably higher than China’s (Fig 3).

**Fig 3.**
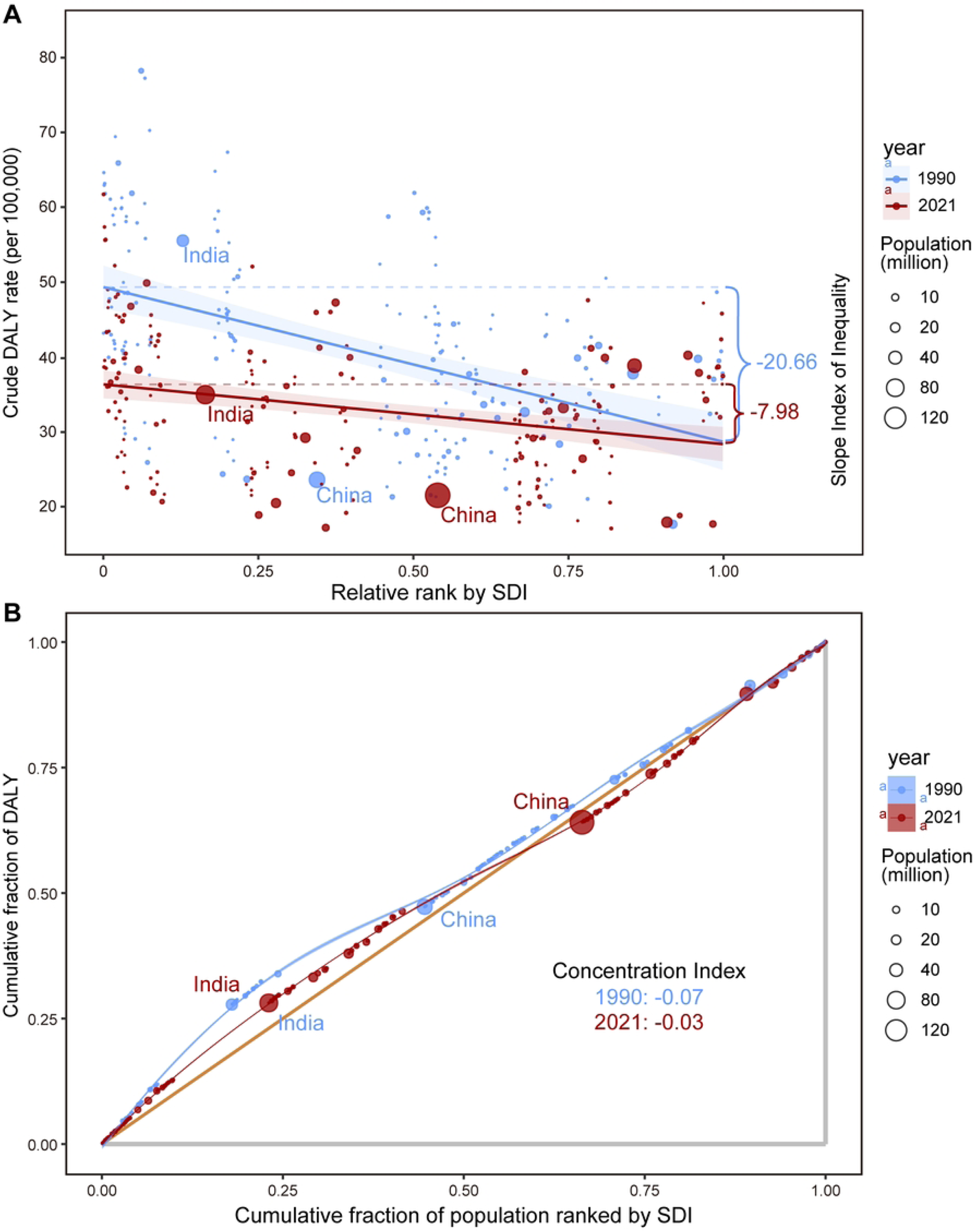
(A) SII analysis: Absolute income-related healthy inequality in POP burden, 1990 vs. 2021. (B) CI analysis: Relative income-related healthy inequality in POP burden, 1990 vs. 2021. SII: slope index of inequality; CI: concentration index; SDI, sociodemographic index.

### Frontier analysis of POP burden

We utilized DALYs data and the SDI for POP across different global regions in 2021 to calculate the effective differences between countries and the frontier. Bolivia, Iceland, Israel, Mexico, and Norway had the largest effective differences (range: 29.77–33.28). Conversely, Thailand, North Korea, Somalia, Japan, and Viet Nam had the lowest DALYs rates for POP, exhibiting the smallest effective differences (range: 0.00–0.21). Significant effective differences suggest potential unrealized health benefits or improvement opportunities (the potential reduction space for global POP-related DALYs) based on a country’s developmental stage. In Fig 4, the frontier is represented by the solid black line, and countries are denoted by dots. Blue dots indicate an upward trend in DALYs, while red dots signify a downward trend. Statistical analysis (S2 Fig) revealed a statistically significant but weak negative correlation between SDI and effective differences (Spearman’s ρ = -0.034, p = 0.0062), with region-specific heterogeneity across SDI tiers (Fig 4 and S2 Table).

**Fig 4.**
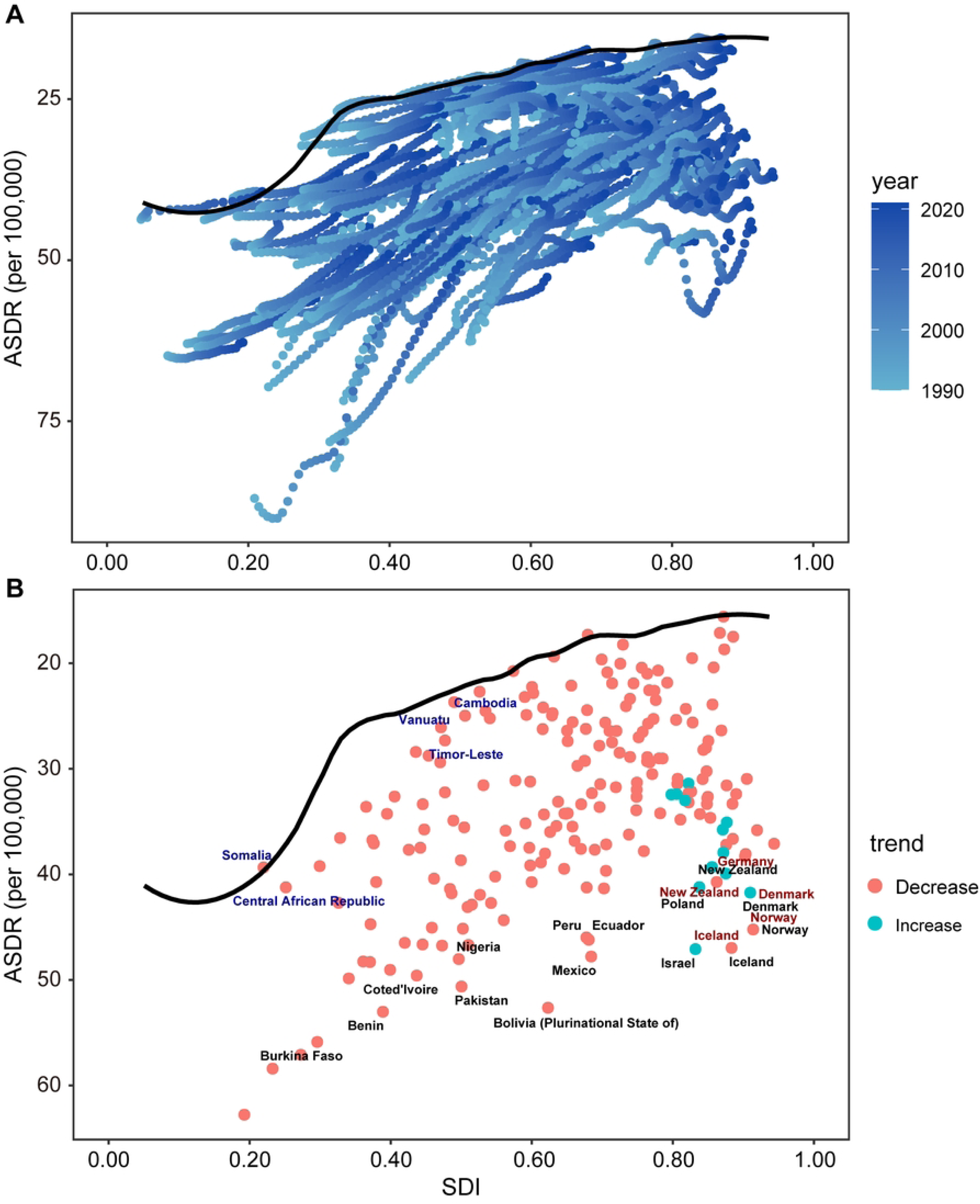
(A) Frontier analysis based on SDI and POP ASDR from 1990 to 2021. (B) Frontier analysis based on SDI and POP ASDR in 2021. ASDR, age-standardized DALY rate; SDI, sociodemographic index.

### APC analysis

Fig 5 illustrates the annual percentage change in POP incidence rates among PMW. The overall net drift was significantly positive in high-middle-SDI regions (0.292%; 95% CI: 0.213% to 0.372%). Conversely, significant negative overall net drifts were observed in middle-SDI regions (−0.190%; 95% CI: -0.262% to -0.117%), low-middle-SDI regions (−0.835%; 95% CI: -0.948% to -0.723%), and low-SDI regions (−0.614%; 95% CI: -0.718% to -0.510%). Globally, the overall net drift was a slight decreasing trend (−0.063%; 95% CI: -0.143% to 0.017%; p = 0.122), although this was not statistically significant. Similarly, the high-SDI regions exhibited a nonsignificant slight increasing trend (0.065%; 95% CI: -0.046% to 0.176%; p = 0.254). The local drifts in the global incidence of POP were predominantly negative across most age groups, indicating declining rates except among the oldest-old (85+ years). Similar downward trends occurred in high-middle and middle-SDI regions. Notably, high-SDI regions demonstrated positive local drifts in the 65–80 age group, reflecting a marked increase in POP incidence. Conversely, the 85+ group in these regions exhibited declining rates. Both low-middle and low-SDI regions showed consistent downward trends in POP incidence among PMW (Fig 5 and S3 Table).

**Fig 5.**
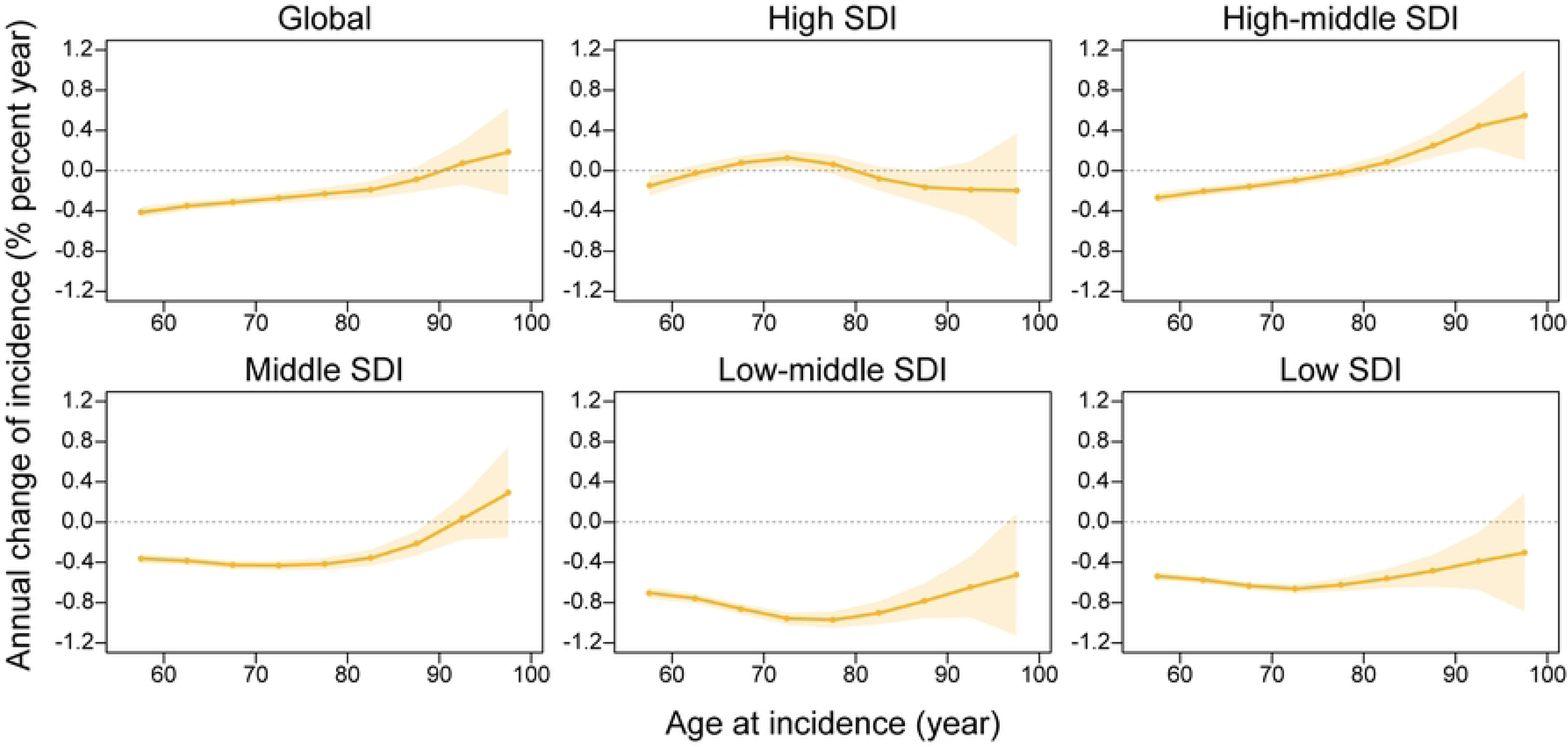
The local drifts of POP incidence rate, 1990–2021.

Fig 6 illustrates the effects of age, period, and birth cohort on POP incidence among PMW. A consistent pattern was observed globally and across SDI regions: incidence rates generally peaked around 70 years of age, declined temporarily, and subsequently increased with advancing age, ultimately reaching a second peak at 90+ years. Among all regions, the middle SDI region demonstrated the steepest age-dependent increase in overall incidence rates, indicating a pronounced age effect. Notably, high-SDI regions exhibited higher incidence rates than the global average in the 65–75 years age group, while low-middle and low-SDI regions showed elevated rates in the 55–70 years age group compared to more developed regions. Period trends reached an equilibrium point (RR = 1) during 2005–2010 globally. After 2010, a rising trend emerged in global RR (RR > 1), with high SDI and high-middle-SDI regions displaying similar trajectories. Importantly, high-middle-SDI regions exerted the strongest period effect on POP incidence among PMW. Conversely, low-middle- and low-SDI regions exhibited declining trends, with RR < 1 after 2010. For birth cohort effects, global RR declined to < 1 for cohorts born after 1930. This pattern was consistent across all SDI regions except high-SDI regions. The steepest decline occurred in low-middle- and low-SDI regions. Notably, women born between 1935 and 1950 in high-SDI regions faced a progressively increasing disease burden, although this burden moderated for subsequent birth cohorts.

**Fig 6.**
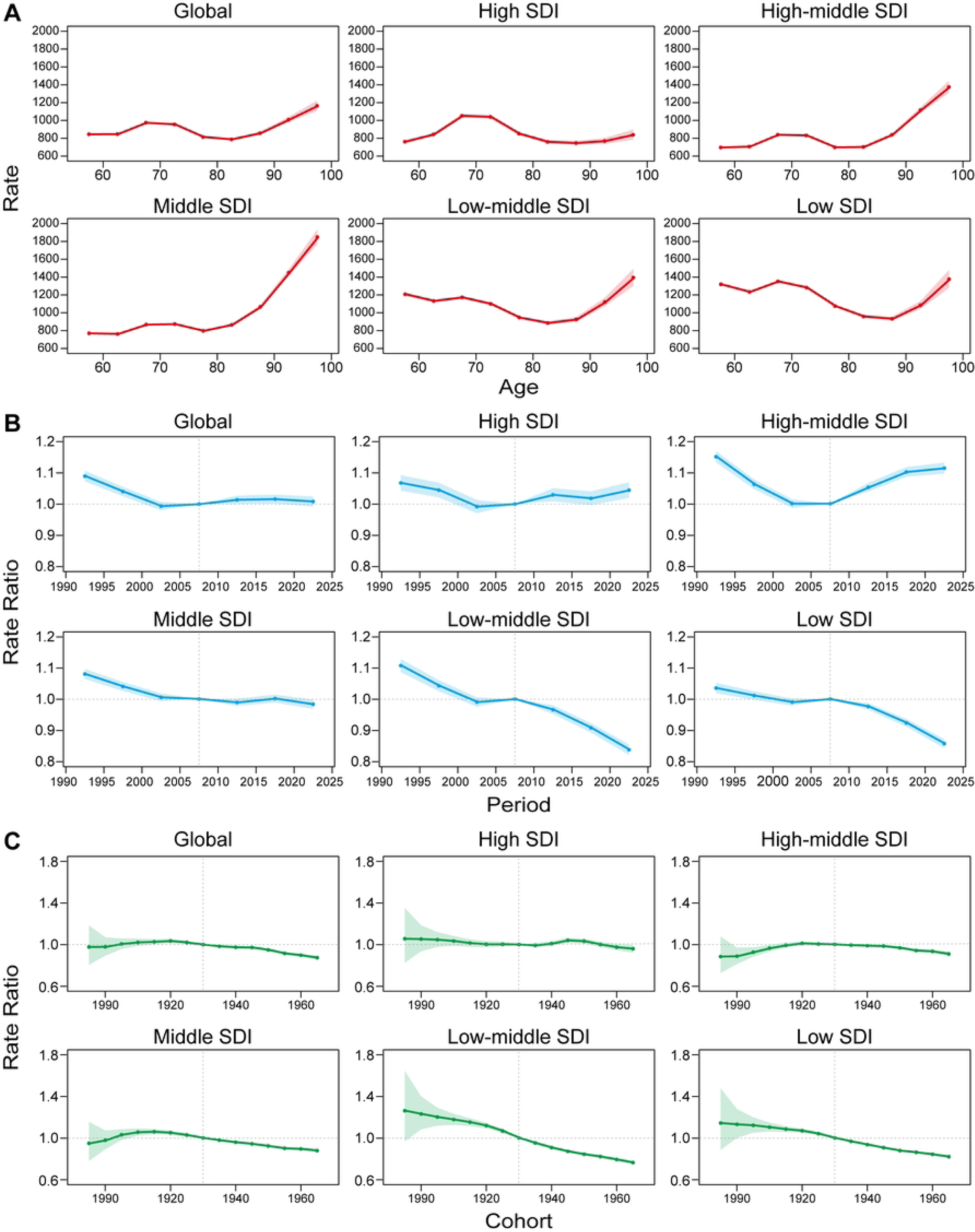
Age, period and cohort effects on POP incidence.

### BAPC analysis

BAPC models were utilized to project that the trends of ASIR, ASPR, and ASDR for POP among PMW will decline between 2022 and 2050. The global prevalence of POP among PMW is expected to reach 114.67 million by 2050, representing a 50.02% increase from 2021. The results indicate that the ASIR is projected to decrease from 816.74 per 100,000 in 2021 to 788.26 per 100,000 in 2050, representing a decline of approximately 3.49%. Similarly, the ASPR is expected to decline from 9912.95 per 100,000 in 2021 to 7961.21 per 100,000 in 2050, reflecting a reduction of approximately 18.04%. Furthermore, the ASDR is projected to decline from 30.20 per 100,000 in 2021 to 23.35 per 100,000 in 2050, a decrease of approximately 22.68%. The shaded area in the figure indicates the potential fluctuation of ASR (Fig 7 and S4 Table).

**Fig 7.**
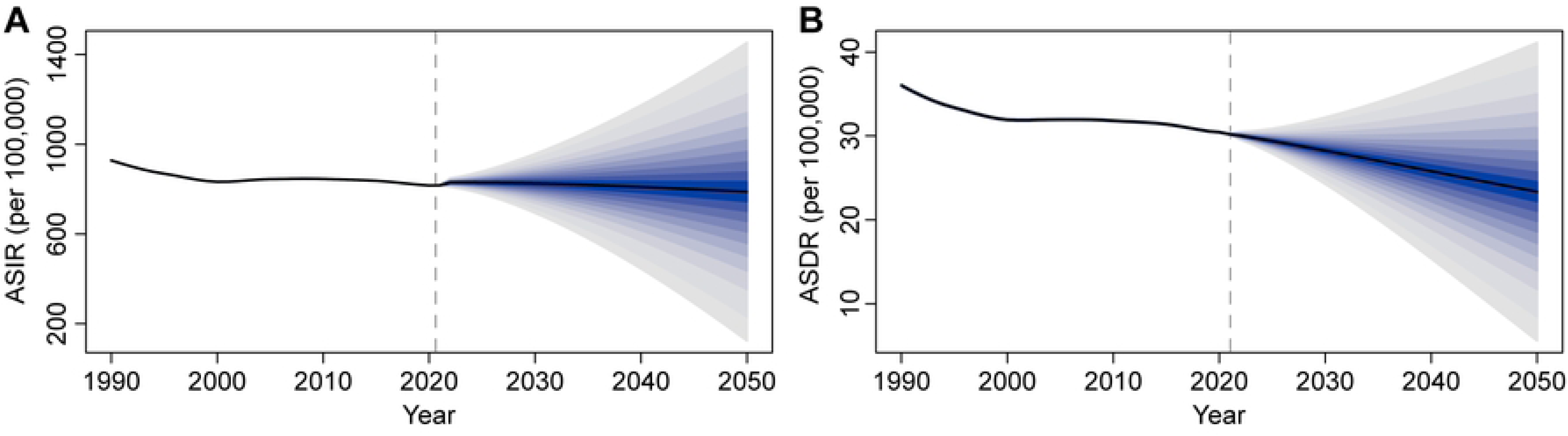
Projected trends in POP among postmenopausal women, 2022–2050: (A) ASIR; (B) ASDR. ASIR, age-standardized incidence rate; ASDR, age-standardized DALY rate.

### Primary MR analysis results for POP risk factors

Based on potential mediators identified in prior studies ^[12–17]^, we conducted UVMR analyses to assess their associations with POP (Fig 8). Complete datasets along with scatter plots, funnel plots, and leave-one-out plots for all results are presented in S6 Table, and S3-S5 Figs. The analysis revealed significant associations between obesity-related anthropometric indicators and POP risk: body mass index (BMI) (OR = 1.124, 95% CI = 1.016–1.243), waist circumference (OR = 1.209, 95% CI = 1.041–1.405), hip circumference (OR = 1.313, 95% CI = 1.056–1.633), body fat percentage (OR = 1.359, 95% CI = 1.023–1.806), whole body fat mass (OR = 1.138, 95% CI = 1.006–1.287), trunk fat percentage (OR = 1.174, 95% CI = 1.017–1.354).

**Fig 8.**
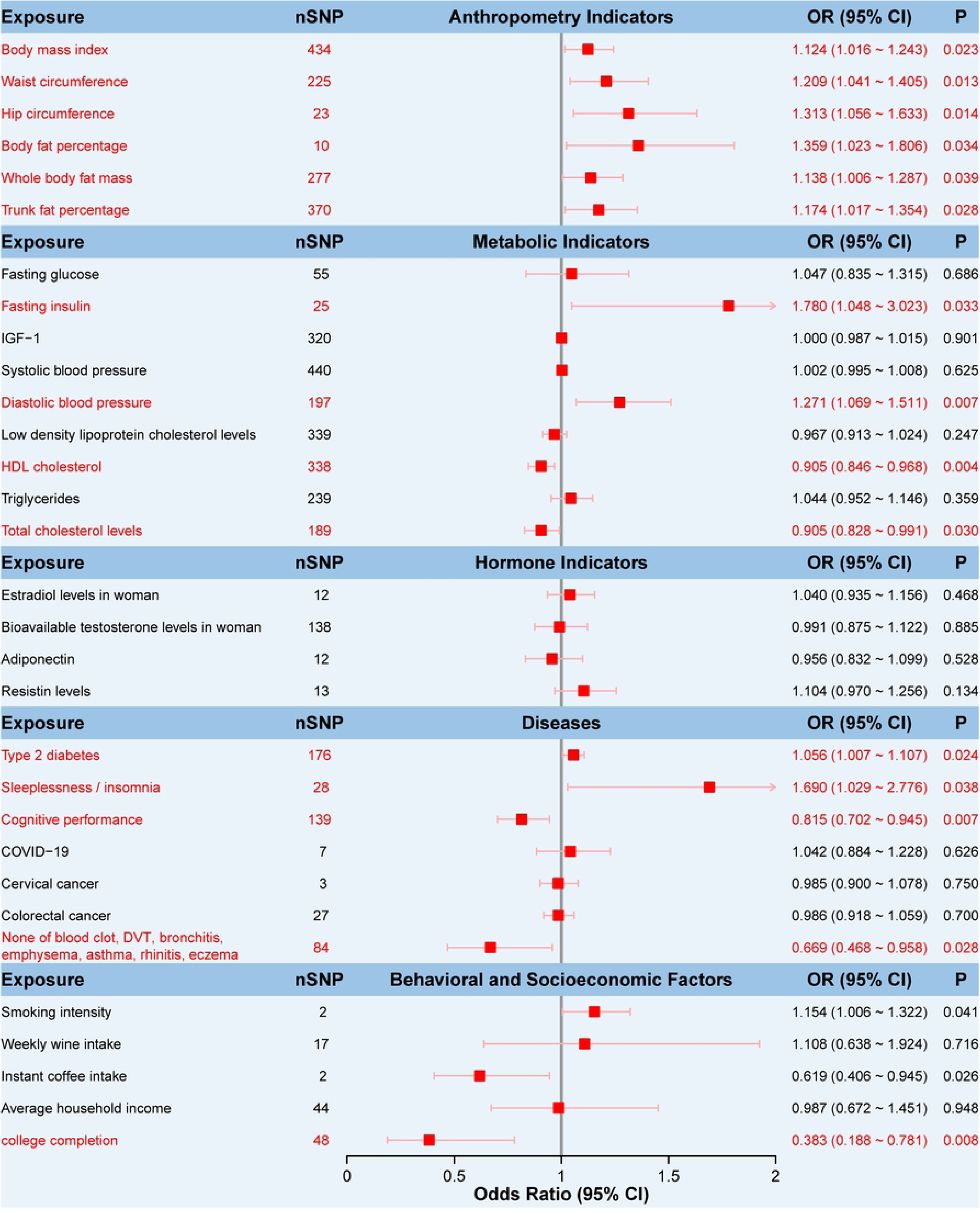
Forest plot of Mendelian randomization analysis. OR, odds ratio.

Among the metabolic indicators analyzed, fasting insulin (OR = 1.78, 95% CI = 1.048–3.023) and diastolic blood pressure (OR = 1.271, 95% CI = 1.069–1.511) were identified as significant risk factors for POP. Conversely, HDL cholesterol (OR = 0.905, 95% CI = 0.846–0.968) and total cholesterol levels (OR = 0.905, 95% CI = 0.828–0.991) demonstrated protective effects against POP development.

Among disease-related factors, type 2 diabetes (OR = 1.056, 95% CI = 1.007– 1.107) and insomnia (OR = 1.690, 95% CI = 1.029–2.776) were identified as risk factors, while cognitive performance showed protective effects (OR = 0.815, 95% CI = 0.702–0.945). Furthermore, the absence of physician-diagnosed conditions, including blood clots, deep vein thrombosis (DVT), bronchitis, emphysema, asthma, rhinitis, and eczema (OR = 0.669, 95% CI = 0.468–0.958), was associated with a reduced risk of POP.

Additionally, college completion (OR=0.383, 95% CI = 0.188–0.781) demonstrated a stronger protective association, consistent with the lower prevalence of POP in high-SDI regions and among populations with higher cognitive levels.

## Discussion

This study, utilizing the latest GBD 2021 data, presents the first systematic integration of the health inequality model, frontier analysis, APC analysis, BAPC prediction model, and MR analysis. It comprehensively analyzes the global epidemiological characteristics of POP among PMW and the evolution of disease burden across multiple dimensions. By comparing data from 1990 to 2021 and projecting trends up to 2050, the study reveals the spatiotemporal heterogeneity of POP, socio-economic driving factors, and the challenges of prevention and control in the context of aging. These findings provide critical evidence-based insights for global health policy formulation. Although the global ASIR, ASPR, and ASDR exhibited declines from 1990 to 2021, indicating varying degrees of progress in the prevention and control of POP across countries, these improvements are strongly associated with major breakthroughs in pelvic floor rehabilitation and advancements in diagnostic and treatment systems during the last 31 years ^[18]^. However, despite these declines, the absolute numbers of incidence, prevalence, and DALYs globally increased by 92.96%, 85.21%, and 83.78%, respectively. Furthermore, high-income countries continue to experience an increasing burden of POP among PMW. These trends demonstrate that population aging and demographic expansion remain the primary drivers of the rising absolute burden.

Stratified analysis of the SDI reveals that the global POP burden among PMW is significantly greater in low-SDI regions. For instance, the ASDR in Niger is 62.77 per 100,000, while in high SDI countries such as Japan, it is only 15.59 per 100,000. A systematic review indicates that the prevalence of stage II-IV POP reaches 58.4% in low SDI countries, compared to only 20.35% in high SDI countries ^[1]^. This disparity may result from the limited access to prenatal care and pelvic floor rehabilitation in low-SDI regions, which often results in delayed intervention and disease progression ^[19]^. Furthermore, factors such as higher rates of vaginal deliveries, lack of professional midwifery services, premature return to manual labor, and engagement in heavy physical work have been identified as significant contributors to the occurrence and progression of POP in low SDI countries ^[20]^. POP is often regarded as a “silent epidemic” in low-SDI regions. Due to social taboos and economic constraints, the rate of medical consultation among affected women remains low, leading to a higher proportion of severe prolapse cases ^[21]^. However, the burden of POP among PMW is increasing in some developed regions. For instance, in Eastern and Central Europe, the EAPC for ASDR is remarkably high (0.723 and 0.307, respectively), primarily driven by rapid population aging. Furthermore, countries such as Poland, Greece, and Japan face critically low fertility rates. As reported by the World Economic Forum, their working-age populations are projected to decline by 35% or more by 2060 ^[22]^, compounding the challenges posed by demographic aging.

According to the health inequality analysis, the SII improved from -20.66 in 1990 to -7.98 in 2021. This trend highlights a reduction in the gap of POP burden among PMW between high and low-SDI regions over the past 31 years from a health equity perspective. However, the burden remains higher in vulnerable groups than in dominant groups. The CI reveals that POP burden is unevenly distributed in low SDI populations, underscoring structural inequalities in health resource allocation. For example, in India, POP incidence among elderly women exhibits a steady upward trend, associated with multiparity and chronic undernutrition ^[23]^. These findings align with MR analysis results, which suggest that elevated total cholesterol levels (potentially reflective of nutritional status in specific populations, e.g., chronic disease or malnutrition ^[24]^) may be associated with reduced risk of POP.

Frontier analysis reveals significant disparities in the burden of POP among PMW across countries stratified by SDI. Notably, the correlation between SDI level and the effective differences is weak, indicating that specific opportunities for improvement exist irrespective of national income level. APC analysis further demonstrates that while PMW in lower-SDI regions exhibit a more pronounced declining trend in POP incidence, the overall incidence rates remain significantly higher than the rates in high-SDI regions. Conversely, although PMW in higher-SDI regions have lower incidence rates across all age groups, these rates exhibit a growth trend. Consequently, low-SDI regions should prioritize enhancing population health literacy and strengthening primary care infrastructure, whereas high SDI counterparts must address aging-driven challenges through optimized chronic disease management (e.g., obesity, respiratory conditions) and mitigation of demographic transition risks. Furthermore, standardizing diagnostic protocols using tools like the POP-Q scale remains essential for improving cross-regional epidemiological data comparability [25].

The prevalence of POP increases with age, leading to a growing clinical demand for effective treatment strategies ^[26]^. The number of surgical procedures for POP in individuals aged over 75 in England rose from 1.5 million in 2006–2007 to 2.5 million in 2014–2015 ^[27]^. Abdominal sacrocolpopexy is regarded as the gold standard treatment for POP. Laparoscopic and robot-assisted surgeries offer better anatomical outcomes, reduced estimated blood loss, and lower overall postoperative complications ^[28]^. Although pelvic floor reconstruction surgery is widely utilized, colpocleisis, with its minimal trauma, is a more suitable option for patients who do not require vaginal intercourse, decline mesh implantation surgery, and prioritize a low risk of complications ^[29]^. A retrospective study revealed that the surgical rate for POP declined from 1987 to 2009. Based on APC analysis, it was determined that the reduction in instrumental vaginal deliveries and obstetric trauma may be significant factors contributing to the decline in the surgical volume of POP ^[30]^. Future research and innovation should focus on optimizing minimally invasive techniques, enhancing repair materials, advancing the understanding of tissue biomechanics, and integrating comprehensive pelvic floor treatments to improve surgical outcomes ^[31]^. Currently, there is no definitive POP prevention strategy; however, potential measures include weight loss, minimizing heavy physical labor, alleviating constipation, optimizing delivery methods, and implementing pelvic floor muscle physical therapy ^[26, 32]^.

The BAPC model predicts that by 2050, the number of POP cases among PMW worldwide will increase to approximately 114.67 million, representing a 50.02% rise from 2021 levels. Although the ASPR is projected to decline to 7961.21 per 100,000 (a reduction of 18.04%), yet the ASIR is expected to decrease by only 3.49% by 2050, with no significant improvement anticipated in the near future. These apparent contradictions suggest that, despite global aging, the increasing adoption of laparoscopic surgery, vaginal reconstruction surgery, and the enhanced utilization of vaginal braces and pelvic floor muscle training have mitigated disease progression ^[33]^. Although the decline in ASPR and ASDR for POP among PMW globally reflects the combined effects of advancements in medical technology, the widespread implementation of preventive measures, and socioeconomic development, the projected increase in the number of patients will still result in a substantial absolute burden and persistent regional disparities. This prediction model offers valuable insights for health policymakers. Moving forward, priority should be given to resource allocation in regions with low SDI and enhanced health management strategies in aging populations, aiming to achieve a more equitable reduction in the burden of disease.

Our MR analyses confirm and quantify the significant causal impact of overall and abdominal obesity on POP risk. Elevated body mass index (BMI), waist circumference, hip circumference, body fat percentage, whole-body fat mass, and trunk fat percentage all demonstrated consistent positive associations. This aligns with substantial prior observational evidence by demonstrating that persistent increases in intra-abdominal pressure and consequent pelvic neurovascular and ligamentous damage induced by abdominal fat accumulation contribute significantly to the observed association between obesity and POP ^[34]^.

Among metabolic traits, fasting insulin and diastolic blood pressure emerged as significant causal risk factors. This suggests that insulin resistance and potentially associated hyperglycemia or vascular dysfunction, alongside elevated diastolic pressure contributing to chronic pelvic floor stress, are important etiological pathways. Conversely, higher levels of both HDL and total cholesterol exhibited significant protective effects, possibly mediated by inhibiting degradation and/or aberrant synthesis of elastin fibers and collagen in pelvic floor connective tissue remodeling, thereby reducing POP incidence ^[35]^. Our MR analysis revealed no significant causal association between estradiol and POP risk (OR = 1.040, 95% CI = 0.935–1.156, P = 0.468). Existing studies indicate that estrogen may influence pelvic floor tissues by regulating collagen metabolism and matrix metalloproteinase (MMP) expression ^[36, 37]^. However, most current evidence derives from small-scale or low-quality studies, with a lack of large-scale randomized controlled trials (RCTs) definitively supporting the therapeutic efficacy of estrogen for POP ^[15]^. Further high-quality studies are needed to clarify its mechanistic role and clinical applicability.

Among disease-related factors, genetically predicted type 2 diabetes mellitus (T2DM) and insomnia were causally linked to increased POP risk. Notably, observational studies suggest that hypertension alone or T2DM alone may not significantly elevate prolapse risk, but their comorbidity synergistically amplifies the risk by approximately two-fold ^[14]^. The novel identification of insomnia as a risk factor suggests potential contributions from chronic stress, impaired healing, hormonal fluctuations, or unmeasured mediators affecting pelvic floor tissues. Higher cognitive performance demonstrated a significant protective effect, reinforcing the importance of neurological factors and potentially reflecting cognitive reserve influencing symptom perception, reporting, healthcare-seeking behavior, or even neuromuscular control mechanisms. Therefore, targeted education on weight control and lipid management for women with lower educational attainment may hold clinical significance for reducing POP risk.

In terms of disease phenotype definition, POP is a clinically heterogeneous syndrome that includes distinct anatomical subtypes such as uterine prolapse, cystocele, and rectocele. Although this study utilized GWAS data on uterine prolapse from the IEU database (2021 release) for Mendelian randomization analysis, the phenotypic heterogeneity across subtypes could affect the validity of inferences regarding POP’s overall genetic architecture. Additionally, although the GBD database contains global population case data, it is challenging to account for systematic differences in data collection and coding, as well as inconsistencies in data source quality within this analytical model. Second, regarding population representativeness, the GWAS data used in this study were primarily derived from European-ancestry populations. Although this helps control for population stratification bias, it may limit the generalizability of findings to other ethnic groups. Finally, in the analysis of exposure factors, the instrumental variables for smoking and coffee intake included only two nonsynonymous SNPs, which may compromise the reliability of causal effect estimates. Future studies should incorporate larger-scale GWAS data to enhance the robustness of these findings.

## Conclusion

This study represents the first systematic assessment of the global burden of POP among PMW, demonstrating that although the overall global burden exhibited a declining trend from 1990 to 2021, significant disparities persist across SDI regions. Specifically, high-SDI regions experienced less pronounced burden reduction due to population aging and lifestyle changes, whereas low-SDI regions exhibited a downward trend but maintained a severe absolute burden. To address these challenges, targeted interventions based on disease burden mapping are recommended: strengthen healthcare infrastructure in low-SDI regions to improve early screening and surgical capacity; enhance health management for elderly PMW in high-SDI regions through integrated pelvic floor rehabilitation programs. These strategies ultimately aim to achieve equitable reductions in POP burden while advancing global health justice.

## Abbreviations

POP: pelvic organ prolapse
GBD: global burden of disease
PMW: postmenopausal women
MR: Mendelian randomization
DALYs: disability-adjusted life years
ASIR: age-standardized incidence rate
ASPR: age-standardized prevalence rate
ASDR: age-standardized DALY rate
SDI: socio-demographic index
SII: slope index of inequality
CI: concentration index
APC: Age-Period-Cohort
BAPC: Bayesian age-period-cohort
EAPC: estimated annual percentage change
UI: uncertainty interval

## Data Availability

All data used in this study were obtained from the GBD 2021 database (https://vizhub.healthdata.org/gbd-results/) and the MRC Integrative Epidemiology Unit (IEU) OpenGWAS project (https://gwas.mrcieu.ac.uk/).

https://gwas.mrcieu.ac.uk/

## Acknowledgements

We sincerely acknowledge the contributions of database researchers whose meticulously developed resources have advanced progress in public health. Their sustained efforts to improve global health data represent an indispensable asset to the scientific community.

## Ethics declarations

### Ethics approval and consent to participate

This is a retrospective, observational cohort study utilizing publicly available database resources; therefore, the requirement for informed consent was waived. The consent was not applicable.

### Consent for publication

All authors have read and agreed to the published version of the manuscript.

## Supporting information

**S1 Fig. ASIR, ASPR, and ASDR of POP in postmenopausal women for the 21 GBD regions by SDI, 1990-2021.**

**S2 Fig. Correlation between SDI and effective differences. S3 Fig. Scatter plots of Mendelian randomization analysis. S4 Fig. Funnel plots of Mendelian randomization analysis.**

**S5 Fig. Leave-one-out plots of Mendelian randomization analysis.**

**S1 Table. ASIR, ASPR, ASDR, and corresponding EAPC of POP in postmenopausal women globally and regions in 1990 and 2021.**

**S2 Table. All results of frontier analysis.**

**S3 Table. Detailed data from Age-Period-Cohort analysis. S4 Table. All results of Bayesian age-period-cohort analysis.**

**S5 Table. Detailed data of Mendelian randomization analysis. S6 Table. All results of Mendelian randomization analysis.**

**S1 File. STROBE-MR checklist.**

## Author Contributions

W.S.: Conceptualization, Funding Acquisition, Project Administration, Resources, Supervision, Writing-Review & Editing. Z.W.: Conceptualization, Data Curation, Formal Analysis, Methodology, Software, Visualization, Writing-Original Draft Preparation. Q.W., Y.R., and Y.W.: Writing-Review & Editing.

